# Biologically Informed Prediction of Response to Neoadjuvant Chemotherapy using Routine Clinical Data in Breast Cancer

**DOI:** 10.64898/2026.01.20.26344418

**Authors:** Xinzhi Teng, Yufeng Jiang, William C. Cho, Hongmei Wang, Junjie Ma, Miaoqing Zhao, Xiangjiao Meng, Haonan Xiao, Qingpei Lai, Xinyu Zhang, Chenyang Liu, Zongxi Li, Tian Li, Haoran Xie, Ge Ren, Andy Lai-Yin Cheung, Jing Cai

## Abstract

**Background:** Early and accurate prediction of pathological complete response (pCR) is essential for personalizing neoadjuvant chemotherapy (NACT) in invasive breast cancer. However, most high-performing predictive models rely on costly, multi-modal data that are not routinely available in standard clinical practice.

**Purpose:** To develop and validate Breast Cancer Biological Multi-modal Information Transfer for Response Prediction Model (**BC-BioMIXER**), a biologically informed predictive model that transfers multi-omics–derived knowledge to routine clinical data, enabling accurate prediction of pathological complete response prior to neoadjuvant chemotherapy initiation.

**Material and Methods:** BC-BioMIXER was developed in a multi-modality cohort of 648 patients with invasive breast cancer (T2–4, any N, M0) incorporating transcriptomic, proteomic, MRI, and clinical data. The model was externally validated in three independent cohorts (total N = 830), including one multi-modality cohort, one clinical trial cohort, and one contemporary real-world cohort. All patients received NACT followed by surgery. The framework employs a teacher–student knowledge-transfer paradigm in which a multi-omics teacher model learns biologically integrated representations that are subsequently transferred to a student model using only routine clinical data. Predictive performance for pCR was benchmarked against a multi-modality reference model and evaluated across cohorts, receptor-defined subgroups (HER2 and hormone receptor [HR]), and treatment groups (NACT with or without immune checkpoint inhibitors [ICI]). Prognostic value was assessed using distant recurrence-free survival (DRFS). The potential to inform immunotherapy decision-making was explored by comparing DRFS between NACT + ICI and NACT-alone groups within model-predicted pCR and non-pCR subgroups.

**Results:** BC-BioMIXER achieved pCR prediction performance comparable to the multi-modality benchmark (AUC 0.82 vs. 0.85; p = 0.271) and demonstrated consistent discrimination across all validation cohorts (AUCs 0.82, 0.81, and 0.80; all p < 0.001). Patients predicted to achieve pCR experienced significantly improved 3-year DRFS (HR = 0.36; 95% CI, 0.20–0.67; p < 0.001). In patients treated with NACT + ICI, BC-BioMIXER showed numerically superior pCR prediction compared with PD-L1 expression alone (AUC 0.84 vs. 0.72; p = 0.08). Notably, within the model-predicted non-pCR subgroup, patients receiving NACT + ICI had significantly inferior DRFS compared with those receiving NACT alone (HR = 2.70; p = 0.032), whereas no significant difference was observed in the predicted pCR subgroup.

**Conclusion:** BC-BioMIXER translates multi-omics–derived biological knowledge into a robust, routine-data–based predictive tool for breast cancer NACT. Its consistent validation across evolving clinical settings and its potential to inform personalized immunotherapy strategies highlight a step toward scalable and accessible precision oncology.

**Highlights:**

1. **Brings multi-omics power to routine clinical practice**: Through cross-modality knowledge transfer, BC-BioMIXER leverages transcriptomic and proteomic data during training to enable highly accurate pCR prediction using only standard MRI and clinical variables (AUC 0.82 vs. 0.85 for full multi-modality benchmark, p=0.271).
2. **Consistently strong and generalizable performance**: Validated in three independent cohorts (total N=830), the model maintained robust pCR discrimination (AUC 0.80–0.82, all p<0.001) across receptor subtypes (HR/HER2) and treatment regimens, including with or without immune checkpoint inhibitors.
3. **Guides personalized immunotherapy de-escalation**: In HER2-negative patients predicted as non-pCR, adding ICI to neoadjuvant chemotherapy was associated with significantly worse distant recurrence-free survival (HR 2.70, p=0.032) compared to chemotherapy alone. This effect was not seen in the predicted pCR group, suggesting the model may help identify patients unlikely to benefit from additional immunotherapy.

## Introduction

Breast cancer is the most prevalent malignancy and a primary driver of cancer-related mortality among women globally (1). For patients presenting with locally advanced or high-risk early-stage disease, neoadjuvant chemotherapy (NACT) serves as a therapeutic cornerstone, facilitating tumor downstaging and expanding eligibility for breast-conserving surgery (2). Despite its utility, clinical response remains highly heterogeneous; while achieving pathological complete response (pCR) is a robust surrogate marker for improved event-free and overall survival, it is currently realized in only 35% to 40% of cases (3). Consequently, the early and accurate prediction of pCR is essential for refining neoadjuvant protocols and advancing the paradigm of precision oncology (4).

Current efforts to optimize NACT increasingly leverage comprehensive molecular profiling to tailor treatment. Recent studies have integrated multi-omics data, incorporating transcriptomics, and proteomics, to refine breast cancer subtyping beyond conventional immunohistochemistry (IHC), facilitating the selection of targeted agents alongside standard anthracycline and taxane-based regimens to improve pCR rates (5–7). Despite the multi-faced and intricate insights provided by multi-omics measurements, their clinical implementation is restricted by high costs, specialized infrastructure, and significant analytical complexity (8).

In routine clinical settings, treatment decisions still rely on IHC-based assessments of hormone receptor (HR) and HER2 status, which often fail to capture the intra-tumoral heterogeneity necessary to predict NACT response accurately. While artificial intelligence (AI) models using medical imaging, such as MRI, show promise in predicting pCR (9,10), those relying solely on radiomic features frequently lack integration with the fundamental biological drivers of chemotherapy resistance (11). This disconnect not only lead to ‘black box’ predictions that lack biological interpretability, but also often result in suboptimal prediction performance (12).

To bridge the gap between the high predictive power of costly multi-omics profiling and the practical limitations of routine clinical feasibility, we introduce BC-BioMIXER, a predictive model that utilizes only routine clinical data while enhancing it with biological insights to deliver more accurate response prediction. Built on a Teacher-Student Knowledge Transfer paradigm (Figure 1b), the model uses an expert “Teacher”, trained on multi-omics data, to transfer its knowledge to a “Student” model trained solely on routine clinical data, guided by both local and global alignment. This approach enriches routine clinical data with multi-omics insights, enabling highly accurate pCR prediction with only low-cost, clinically feasible inputs.

**Figure 1.**
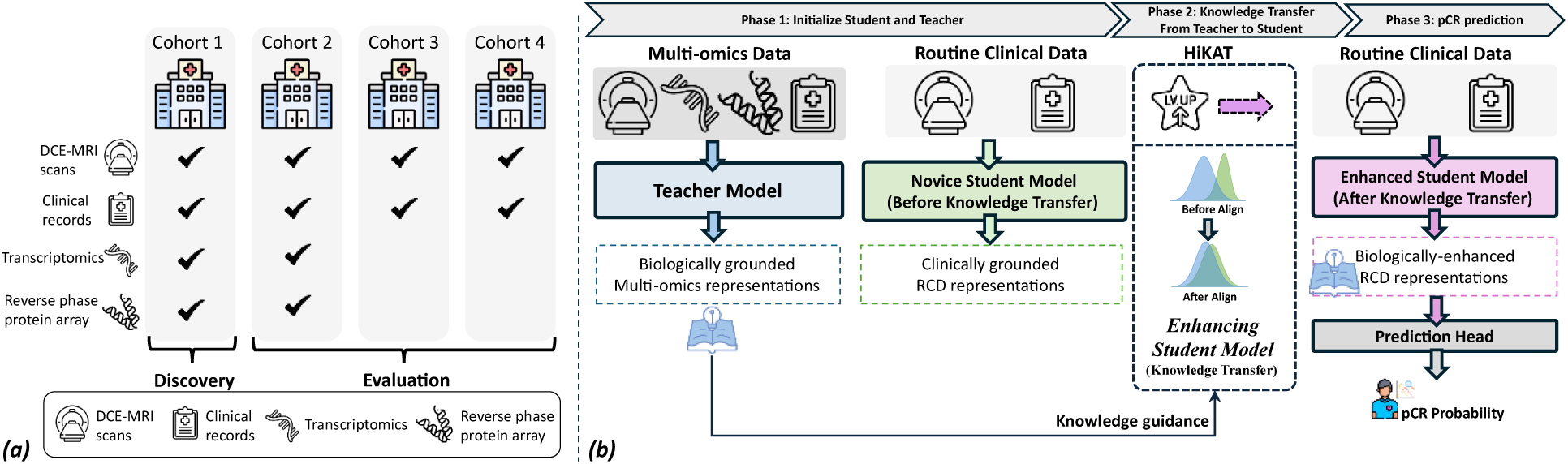
Study overview. (a) Study cohorts and data availability. Four inter-institutional cohorts of invasive breast cancer patients receiving NACT followed by surgery from different countries. Cohorts 1–2 (discovery set) contain comprehensive biological data; Cohorts 3–4 (evaluation set) contain only routine clinical data. (b) Workflow of BC-BioMiXER. The expert Teacher model learns comprehensive molecular wisdom from multi-omics data (Multi-omics Representations), while the novice Student model generates macro-scope knowledge from routine clinical data (RCD Representations). The Hierarchical Knowledge Alignment and Transfer (HiKAT) module aligns local and global representations to transfer biological knowledge to the Student. After knowledge infusion, the enhanced student model enables robust pCR prediction using only routine clinical data at inference.

In this study, we detail the development and rigorous validation of BC-BioMIXER for pCR prediction in patients with locally advanced breast cancer. Performance is comprehensively evaluated across independent cohorts, receptor subtypes (HER2/HR), and regimens incorporating immune checkpoint inhibitors (ICI), underscoring its potential to inform personalized neoadjuvant strategies.

## Methods

### Materials

Four cohorts (Cohorts #1–4) of biopsy-proven invasive breast cancer patients who underwent neoadjuvant chemotherapy followed by surgery were collected as shown in Figure 1; the patient inclusion flowchart is provided in Supplementary Materials S1. All patients received neoadjuvant chemotherapy consisting of anthracycline–cyclophosphamide followed by a taxane (AC→T or T→AC). Patients with HER2-positive status received additional anti-HER2 therapy. A subset of Cohort 2 (HER2-negative tumors) and Cohort 4 (triple-negative tumors) received ICI in addition to standard neoadjuvant chemotherapy. For all cohorts, dynamic contrast-enhanced MRI (DCE-MRI) and clinical variables (HER2 status, HR status, and age at diagnosis) were collected. For Cohorts 1 and 2, transcriptomic (bulk mRNA) expression data (∼19,000 mRNAs), profiled using the Agilent 44K expression array (5), and proteomic data (139 proteins/phosphoproteins), assayed using the GLP28470 Reverse Phase Protein Array (RPPA) platform at George Mason University, were also obtained (7).

### Pathology data

Baseline HER2 and hormone receptor (HR) status were evaluated in core biopsy samples using immunohistochemistry (IHC) or fluorescence in situ hybridization (FISH). HR positivity was defined as ≥ 5% tumor staining, consistent with the protocols of the development cohort. HER2 positivity was defined by an IHC score of 3+ or the presence of FISH amplification. The primary analysis endpoint was the assessment of pCR at surgery, defined as complete tumor disappearance regardless of the presence or absence of ductal carcinoma in situ (ypT0/is, ypN0) (13).

### Medical Image data

Breast MRI data were acquired using standardized 3D gradient-echo (GE) sequences in the axial plane with bilateral coverage. Imaging was performed with an anterior-posterior frequency direction and right-left phase direction. Key acquisition parameters included a field of view (FOV) and matrix size appropriate for complete bilateral coverage, in-plane resolution of 1.0–1.4 mm, and slice thickness of 2.2–2.5 mm without inter-slice gap. Fat suppression was applied, and imaging parameters such as TR, TE, and flip angle were optimized for high-quality dynamic contrast-enhanced imaging. Parallel imaging techniques were used to reduce scan time, with sequence acquisition times ranging from 40 to 100 seconds and total post-contrast imaging durations of at least 5–8 minutes. All patients received gadolinium-based contrast agents at a dose of 0.1 mmol/kg body weight. Detailed MRI acquisition parameters for each cohort are provided in Supplementary Table 1. The radiomics features, in compliant with Image Biomarker Standardisation Initiative (IBSI) were extracted on pre-contrast phase, early contrast phase (∼150s after injection) and late-contrast phase (∼450s after injection) within functional tumor volume (10).

### Transcriptomics and proteomics data

We utilized the I-SPY2-990 Data Resource (5), which provides comprehensive gene expression and protein/phosphoprotein data for Cohorts #1 and #2, as illustrated in Figure 1. For all patients in these cohorts, pre-treatment tumor samples were collected prior to therapy initiation, and full transcriptome profiles covering over 19,000 genes were generated using the Agilent 44K platform. Additionally, normalized Laser Capture Microdissection–Reverse Phase Protein Array (LCM-RPPA) data for 139 key signaling proteins and phosphoproteins were available. Importantly, both the transcriptomic and RPPA data are paired with baseline MRI scans, forming the foundation for cross-modality knowledge transfer, which is central to this study. The I-SPY2-990 Data Resource is publicly accessible via the NCBI Gene Expression Omnibus (Super-Series GSE196096, including Sub-Series GSE194040 [mRNA] and GSE196093 [RPPA]).

### Development of BC-BioMIXER

The development workflow of BC-BioMIXER, as detailed in **Figure 2**, proceeds through three sequential phases. **Phase 1** initializes the dual-stream architecture by establishing an expert Teacher model to encode multi-omics representations and a novice Student model to process routine clinical data. **Phase 2** facilitates knowledge distillation via the HiKAT module, which dynamically aligns molecular and clinical features to infuse biological wisdom into the Student. Finally, **Phase 3** deploys the knowledge-enhanced Student model to independently predict pCR using solely routine clinical inputs at inference.

**Figure 2.**
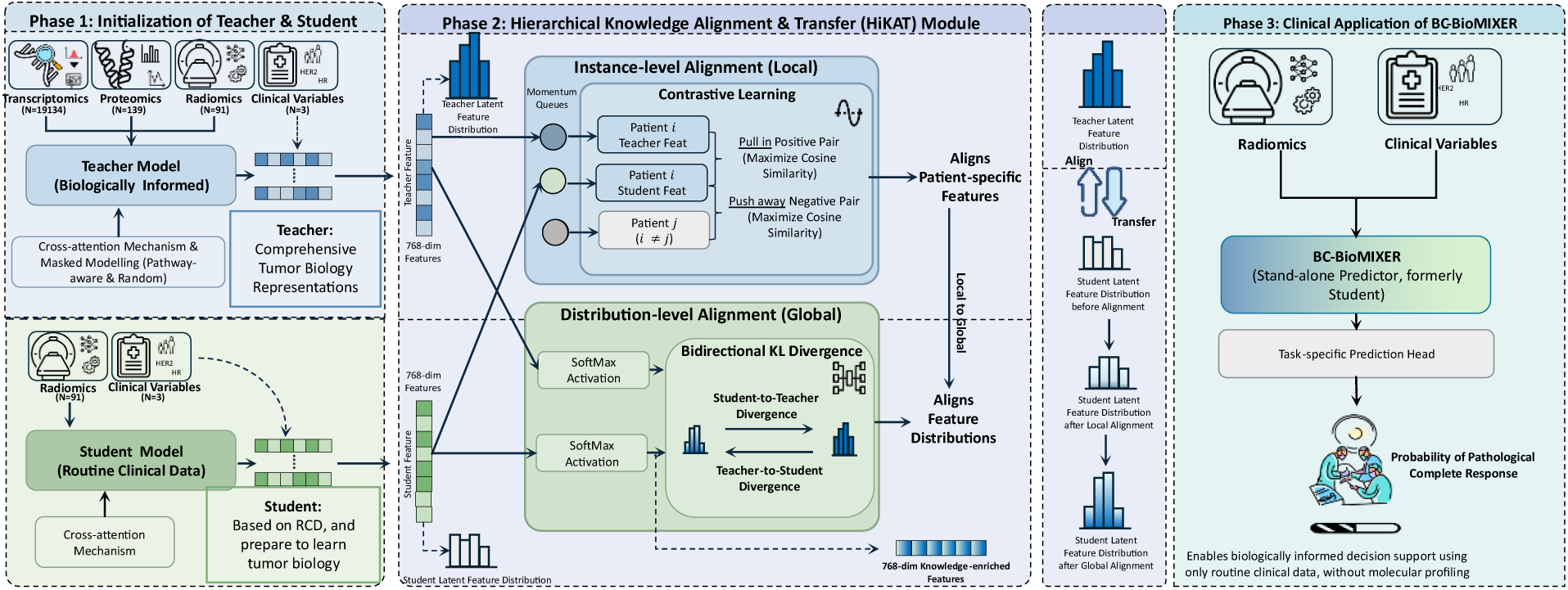
BC-BioMIXER development workflow. The framework consists of three phases. Phase 1: Initialization of Teacher & Student. The teacher model is trained on comprehensive biological data (transcriptomics, proteomics, radiomics, and clinical variables) using cross-attention and masked modeling to learn comprehensive tumor biology representations. The student model is trained on routine clinical data (radiomics and clinical variables) using cross-attention. Phase 2: Hierarchical Knowledge Alignment & Transfer (HiKAT) Module. Knowledge is transferred through two levels: (1) Instance-level alignment (Local) uses contrastive learning to align patient-specific features by pulling positive pairs together and pushing negative pairs apart; (2) Distribution-level alignment (Global) uses bidirectional KL divergence to align overall feature distributions between teacher and student. Phase 3: Clinical Application. The enhanced student model (BC-BioMIXER) serves as a stand-alone predictor using only routine clinical data to generate biologically informed pCR predictions without requiring molecular profiling.

#### Phase 1: Initialization of Student and Teacher

##### Overview

In Phase 1, we initialize two complementary models with distinct roles. The Teacher model serves as a biologically informed reference, designed to capture comprehensive tumor characterization through the integration of multi-omics data. In contrast, the Student model is initialized using routine clinical data (RCD) and is intended to learn clinically accessible imaging representations that can ultimately approximate the molecular knowledge encoded by the Teacher. This phase establishes the biological and clinical foundations required for subsequent cross-modal knowledge transfer.

##### Teacher Model Construction

The Teacher model integrates heterogeneous multi-omics and clinical data to construct a unified representation of tumor biology. Specifically, the input modalities include:

1. 19,134 mRNA transcriptomic features,
2. 139 protein and phosphoprotein measurements derived from reverse-phase protein arrays (RPPA),
3. 91 repeatable and reproducible radiomics features defined according to the IBSI, and
4. key clinical variables, including age at diagnosis, hormone receptor status, and HER2 status.

##### Learning Biologically Meaningful Representations via Masked Modeling

To encourage the Teacher model to learn biologically meaningful and generalizable molecular representations—rather than superficial correlations—we employ a masked modeling strategy during training. Subsets of gene or protein features are masked and subsequently reconstructed by the model, requiring it to infer latent biological structure from the observed data (14). Masking is applied in two complementary ways. First, pathway-aware masking targets predefined gene sets corresponding to known biological processes and tumor-related pathways, including luminal differentiation (15), HER2-enriched signaling (16), immune activity (16–20), proliferation (15,16,21), and DNA damage repair deficiency (22,23), as well as candidate novel molecular signatures (24–26). This strategy promotes the learning of established, domain-specific biological knowledge. Second, random masking is applied to encourage the discovery of broader molecular dependencies and potential novel interactions beyond predefined pathways (27). Together, these masking strategies enable the Teacher model to capture both well-characterized and emerging aspects of tumor biology.

##### Student Model Initialization

The Student model is initialized using routine clinical data that are readily available in standard oncology workflows, specifically the 91 repeatable radiomics features and the same set of clinical variables (age at diagnosis, hormone receptor status, and HER2 status). Similar to the Teacher, the Student incorporates a cross-attention mechanism to model dependencies within its input features, enabling the extraction of structured radiological and clinical representations despite the absence of direct molecular information at this stage.

##### Outcome of Phase 1

Phase 1 establishes a biologically informed Teacher model with comprehensive multi-omics understanding and a clinically grounded Student model initialized to capture RCD-derived tumor characteristics. This complementary initialization provides the necessary foundation for subsequent phases of knowledge transfer, in which molecular insights learned by the Teacher are progressively distilled into clinically accessible imaging representations.

#### Phase 2: Cross-Modal Knowledge Transfer

##### Overview

The objective of Phase 2 is to transfer biologically informed representations learned by the Teacher model to the Student model, which at this stage relies exclusively on routine clinical data (RCD). This transfer is implemented through a Hierarchical Knowledge Alignment and Transfer (HiKAT) module, in which the Student’s 768-dimensional representations are progressively aligned with the corresponding Teacher representations. To ensure both robustness and biological fidelity, knowledge transfer is guided by complementary objectives operating at two hierarchical levels: patient-specific (local) alignment and feature distribution (global) alignment.

##### Instance-Level Alignment (Local)

To facilitate patient-specific knowledge transfer between RCD-derived and molecular-derived representations, we perform instance-level alignment between the Student and Teacher features. A cross-modal contrastive learning framework with momentum queues is adopted to explicitly align representations at the individual patient level. For each patient, the Student (RCD-based) and Teacher (multi-omics–based) features constitute a positive pair, whose cosine similarity is maximized, while features from different patients are treated as negatives and encouraged to separate in the embedding space. This local alignment strategy addresses heterogeneity between clinical imaging data and molecular profiles by enforcing consistency between RCD and molecular representations of the same tumor, while preserving inter-patient separability. As a result, the Student is encouraged to learn patient-specific imaging features that reflect underlying molecular characteristics, supporting stable and biologically meaningful knowledge transfer.

##### Distribution-Level Alignment (Global)

While instance-level alignment enforces correspondence at the patient level, it does not explicitly constrain the overall structure of the learned representations. To capture higher-order consistency between RCD-derived and molecular-derived feature responses, we additionally perform feature distribution alignment at the global level. Specifically, SoftMax normalization is applied to the 768-dimensional embeddings of both the Student and Teacher along the feature dimension, transforming them into probability distributions that encode relative patterns of feature activation. Distributional consistency is enforced by minimizing the bidirectional Kullback–Leibler (KL) divergence between the Student and Teacher distributions, jointly constraining both Student-to-Teacher and Teacher-to-Student divergence. This symmetric formulation is critical for biologically faithful knowledge transfer. By constraining divergence in both directions, the Student is encouraged to preserve the full spectrum of molecular information encoded by the Teacher, rather than learning a simplified or partial approximation. By aligning feature distributions rather than individual feature vectors alone, this global alignment strategy provides a more holistic mechanism for cross-modal knowledge transfer. It enables the Student to capture not only which molecular-related features are reflected in imaging-derived representations, but also their relative activation patterns across the entire feature space, thereby enhancing biological interpretability and clinical relevance.

##### Outcome of Phase 2

Through the complementary use of instance-level and distribution-level alignment objectives, the Student progressively acquires molecular-grade biological insights despite relying solely on imaging and clinical data. This phase enables the distillation of complex multi-omics knowledge into clinically accessible representations, laying the groundwork for downstream translational and prognostic applications.

#### Phase 3: Clinical Application of BC-BioMIXER for pCR Prediction

##### Overview

In Phase 3, we evaluate the final model, termed BC-BioMIXER, in a clinically relevant downstream task: prediction of pCR to therapy. At this stage, the Teacher model is no longer involved. BC-BioMIXER, corresponding to the trained Student model enriched through hierarchical cross-modal knowledge transfer, operates as a stand-alone predictor using only RCD. This phase directly assesses the translational potential of BC-BioMIXER in a real-world oncology setting.

##### pCR Prediction Using BC-BioMIXER

BC-BioMIXER generates patient-level representations from RCD, including repeatable radiomics features and standard clinical variables available at diagnosis. These representations are subsequently passed to a task-specific prediction head to estimate the probability of achieving pCR following treatment. Model training and evaluation are performed in a supervised manner, with pCR as the clinical endpoint. By design, BC-BioMIXER requires no molecular profiling at inference time, reflecting realistic clinical workflows in which comprehensive multi-omics data are often unavailable or impractical. Importantly, the imaging-derived representations used for prediction have been biologically enriched through prior knowledge transfer from a multi-omics–informed Teacher model, enabling BC-BioMIXER to implicitly leverage molecular-level insights while remaining clinically deployable.

##### Clinical Rationale and Translational Significance

Pathological complete response is a well-established surrogate endpoint associated with favorable long-term outcomes in breast cancer. Accurate pre-treatment prediction of pCR may support patient stratification, guide treatment intensification or de-escalation, and inform personalized therapeutic decision-making. By distilling molecular oncology knowledge into an imaging-based model, BC-BioMIXER aims to improve the biological interpretability and predictive performance of pCR estimation while maintaining feasibility for routine clinical use.

### Model evaluation

Model performance was evaluated across cohorts 2, 3, and 4, as illustrated in Figure 3. Multiple complementary analyses were conducted to assess the predictive performance, generalizability, and clinical utility of BC-BioMIXER.

**Figure 3.**
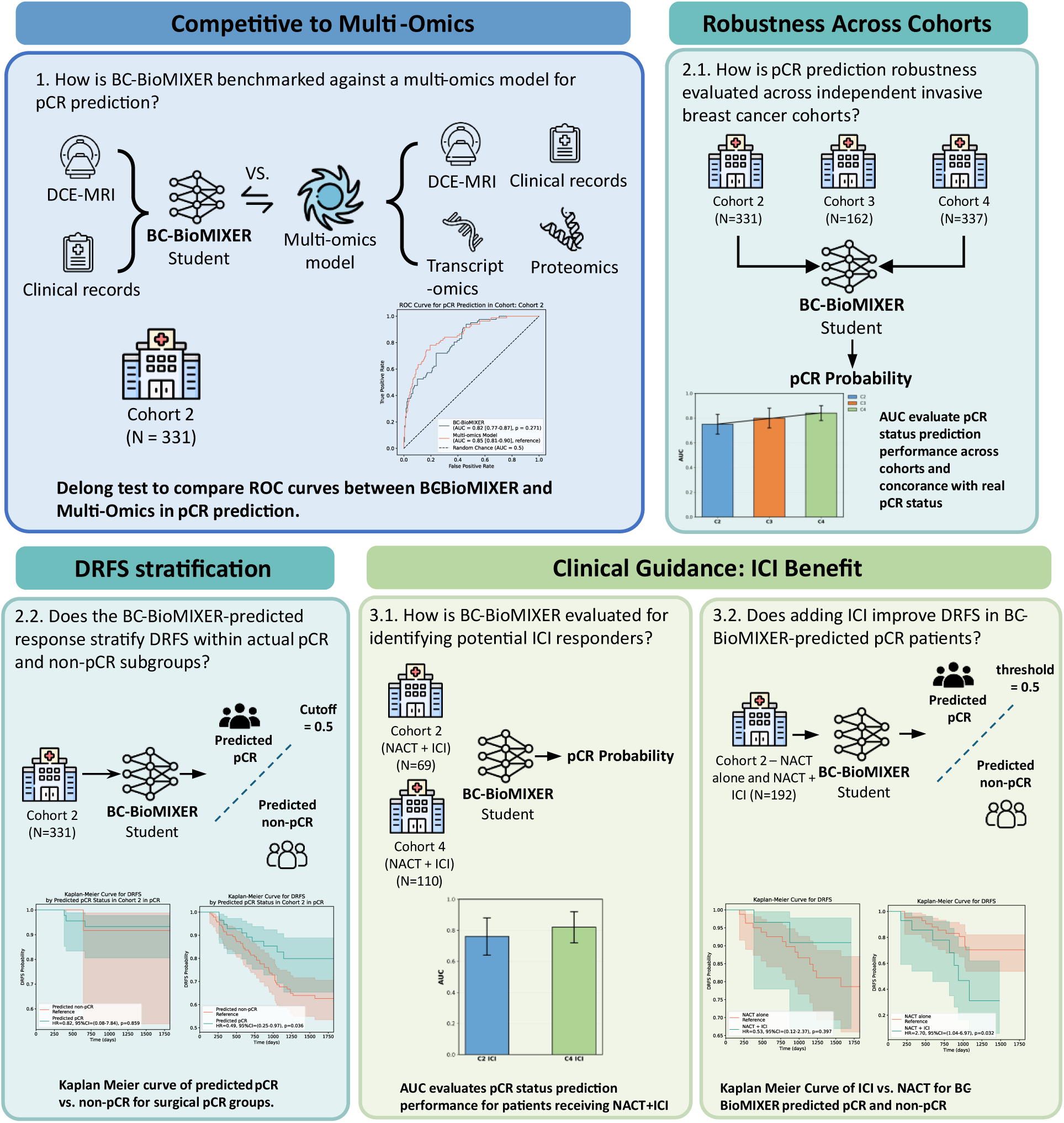
BC-BioMIXER: pCR prediction evaluation and guidance in immunotherapy. Multi-cohort validation of predictive performance and treatment guidance. (1) Competitive to Multi-Omics: BC-BioMIXER (using only DCE-MRI and clinical records) is benchmarked against a multi-omics model (using DCE-MRI, clinical records, transcriptomics, and proteomics) for pCR prediction in Cohort 2 (N=331), with DeLong test comparing ROC curves. (2.1) Robustness Across Cohorts: pCR prediction performance is evaluated across three independent invasive breast cancer cohorts (Cohorts 2–4), assessing AUC and concordance with actual pCR status. (2.2) DRFS Stratification: Kaplan-Meier analysis examines whether BC-BioMIXER-predicted pCR status stratifies distant recurrence-free survival (DRFS) within actual pCR and non-pCR subgroups. (3) Clinical Guidance for ICI Benefit: (3.1) BC-BioMIXER is evaluated for identifying potential immune checkpoint inhibitor (ICI) responders in patients receiving NACT+ICI (Cohort 2, N=69; Cohort 4, N=110); (3.2) Kaplan-Meier analysis assesses whether adding ICI improves DRFS in BC-BioMIXER-predicted pCR versus non-pCR patients (Cohort 2, NACT alone vs. NACT+ICI, N=192).

First, we examined the extent to which the pCR prediction performance of BC-BioMIXER approximates that of a multi-omics reference model. This reference model was constructed using latent representations derived from the Teacher model and trained to predict pCR in cohort 1, serving as an upper-bound benchmark for biologically informed prediction.

Second, we assessed the generalizability of BC-BioMIXER across independent validation cohorts, with particular emphasis on cohort 4, which comprises consecutively collected real-world cases. To explore potential heterogeneity in predictive performance, subgroup analyses were performed according to receptor status.

Third, we investigated whether BC-BioMIXER provides incremental prognostic information beyond the observed pathological complete response status assessed at surgery. Specifically, 3-year distant recurrence-free survival (DRFS) was compared between patients predicted to achieve pCR and those predicted to have residual disease, evaluating the ability of model-derived predictions to further stratify long-term outcomes.

Fourth, we evaluated the performance of BC-BioMIXER in predicting pCR among patients treated with neoadjuvant chemotherapy combined with immune checkpoint inhibitors (NACT + ICI), reflecting contemporary treatment paradigms. The predictive value of BC-BioMIXER was directly compared with that of the established immunotherapy biomarker programmed death-ligand 1 (PD-L1, CD274). To inform therapeutic decision-making, DRFS outcomes were additionally compared between patients receiving NACT + ICI and those treated with NACT alone, stratified by BC-BioMIXER-predicted pCR and non-pCR subgroups.

For all analyses, patients were dichotomized into predicted pCR and predicted non-pCR groups using a predefined probability threshold of 0.5.

### Statistical analysis

The concordance between predicted pCR probability and actual pCR status was evaluated using the area under the receiver operating characteristic curve (AUC-ROC). Comparisons between ROC curves were performed using the DeLong test (28). DRFS was assessed by constructing Kaplan-Meier survival curves, and differences in survival distributions between groups were analyzed using the log-rank test. All statistical analyses were conducted in Python (version 3.10), utilizing key packages including scikit-learn for ROC and AUC calculations (29), lifelines for survival analysis (30), and SciPy for statistical testing (31).

## Results

### Patient Characteristics

A total of 1,478 patients were included across four cohorts: cohort 1 (n = 6 8; mean age, 9 years), cohort 2 (n = 331; mean age, 8 years), cohort 3 (n = 162; mean age, 7 years), and cohort (n = 337; mean age, 49 years). Baseline clinicopathological characteristics are summarized in Table 1. Cohort 4, which represents consecutively collected real-world cases, included a significantly higher proportion of HER2-positive tumors, reflecting institutional practice patterns in which neoadjuvant chemotherapy is preferentially administered to patients with HER2-positive disease. Among patients treated with NACT + ICI, all patients in cohort 2 (n = 69; mean age, 49 years) were HER2-negative, whereas all patients in cohort (n = 110; mean age, 50 years) had triple-negative breast cancer. The median follow-up time for cohort 2 was 3.29 years.

**Table 1.** Patient characteristics of inclusion patients.

| Characteristics | Discovery cohort 1 | Independent test cohort 2 | Independent test cohort 3 | Independent test cohort 4 |
| --- | --- | --- | --- | --- |
| Number of Patients | 648 | 331 | 162 | 337 |
| Age at Screening |  |  |  |  |
| Average (Range) | 49 (24-73) | 48 (23-77) | 48 (28-69) | 49 (24-77) |
| HER2 |  |  |  |  |
| Positive | 169 (23.57%) | 73 (27.86%) | 53 (32.72%) | 180 (53.41%) |
| Negative | 479 (76.43%) | 258 (72.14%) | 109 (67.28%) | 157 (46.59%) |
| HR |  |  |  |  |
| Positive | 342 (53.28%) | 191 (57.63%) | 72 (44.44%) | 157 (46.59%) |
| Negative | 306 (46.72%) | 140 (42.37%) | 90 (55.56%) | 180 (53.41%) |
| Missing |  |  |  |  |
| Menopausal status |  |  |  |  |
| Premenopausal | 317 (48.92%) | 160 (48.33%) | \ | 202 (59.94%) |
| Peri/Postmenopausal | 226 (34.88%) | 105 (31.72%) | \ | 135 (40.06%) |
| Missing | 105 (16.20%) | 66 (19.94%) | \ | 0 (0%) |
| Primary tumor classification |  |  |  |  |
| T2 | \ | \ | \ | 236 (70.03%) |
| T3-T4 | \ | \ | \ | 101 (29.97%) |
| Nodal involvement |  |  |  |  |
| N0 | \ | \ | \ | 62 (18.40%) |
| N1 | \ | \ | \ | 164 (48.67%) |
| N2 | \ | \ | \ | 67 (19.88%) |
| N3 | \ | \ | \ | 44 (13.06%) |
| Receiving additional immunotherapy |  |  |  |  |
| Yes | 0 | 69 (20.85%) | 0 | 110 (32.64%) |
| No | 648 (100%) | 262 (79.15%) | 162 (100%) | 227 (67.36%) |
| Treatment Response at Surgery |  |  |  |  |
| non-pCR | 447 (68.98%) | 180 (68.70%) | 115 (70.99%) | 189 (56.08%) |
| pCR | 201 (31.02%) | 82 (31.30%) | 47 (29.01%) | 148 (43.92%) |

### Benchmarking BC-BioMIXER Against a Multi-Omics Reference Model

To assess the predictive performance of BC-BioMIXER for pCR, we benchmarked it against a multi-omics reference model in cohort 2. BC-BioMIXER achieved an AUC of 0.82 (95% CI, 0.77–0.87), compared with an AUC of 0.85 (95% CI, 0.81–0.90) for the multi-omics model. Although a modest numerical reduction in performance was observed, the difference between the two models was not statistically significant (DeLong’s test, p = 0.271; Figure 4a). These results indicate that BC-BioMIXER achieves pCR prediction performance comparable to that of a substantially more resource-intensive multi-omics approach.

**Figure 4.**
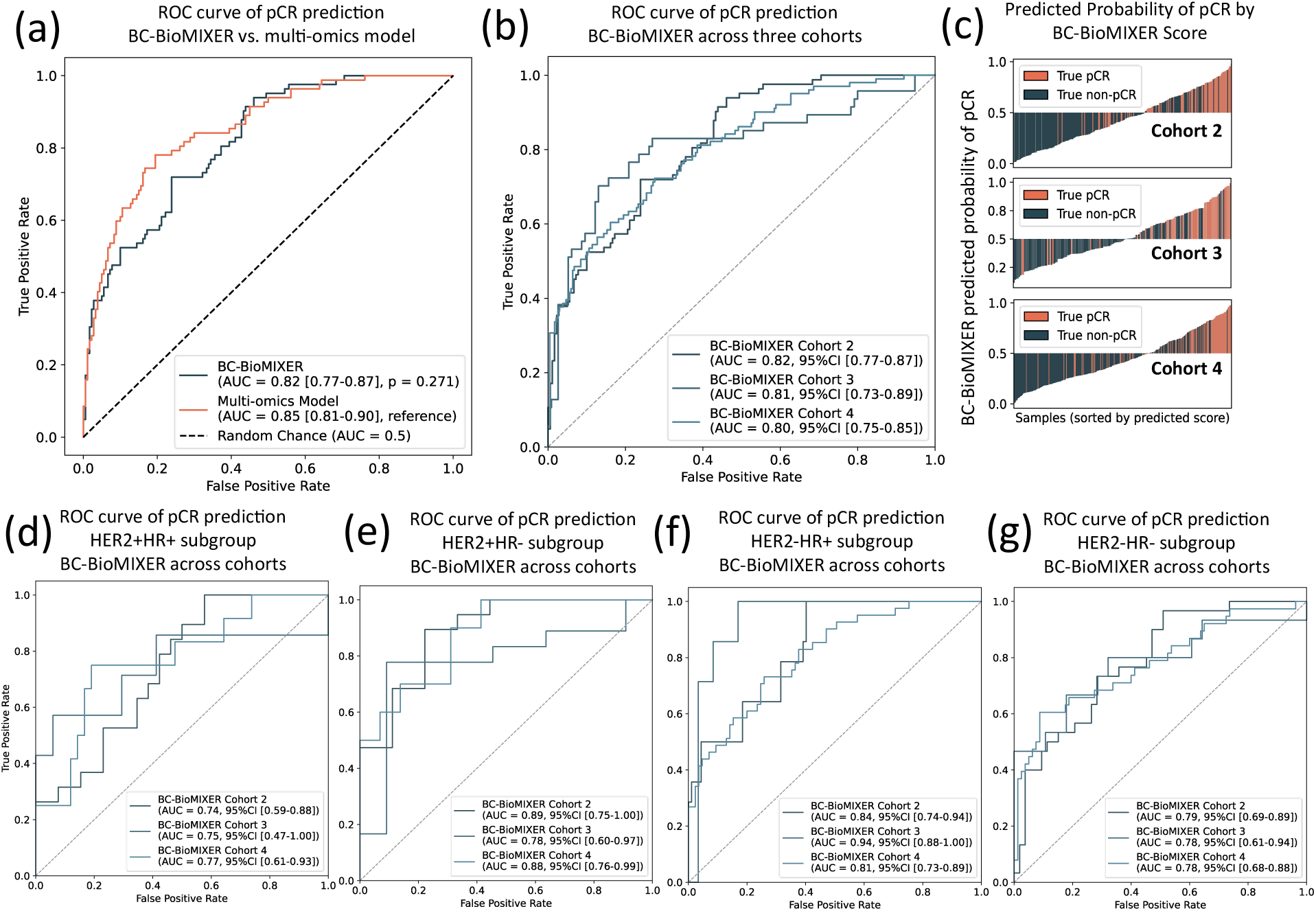
Predictive performance of BC-BioMIXER for pCR. (a) ROC curve comparing BC-BioMIXER with a multi-omics benchmark for pCR prediction in Cohort 2. (b) ROC curves for BC-BioMIXER pCR prediction across three independent cohorts. (c) Distribution of predicted pCR probabilities according to true response status (red: pCR; blue: non-pCR) across Cohorts 2–4. (d–g) ROC curves for pCR prediction within receptor-defined subgroups: (d) HER2+/HR+, (e) HER2+/HR−, (f) HER2−/HR+, and (g) HER2−/HR−.

### Generalizability Across Independent International Cohorts

We next evaluated the generalizability of BC-BioMIXER across three independent international cohorts. As shown in Figure 4b, the model maintained consistent predictive performance, with AUCs of 0.82 (95% CI, 0.77–0.87) in cohort 2, 0.81 (95% CI, 0.73–0.89) in cohort 3, and 0.80 (95% CI, 0.75–0.85) in cohort 4. The distribution of predicted pCR probabilities (Figure 4c) demonstrated clear separation, with patients achieving pCR exhibiting higher predicted probabilities than those with residual disease, supporting robust discrimination across diverse clinical settings.

### Subgroup Performance Stratified by Hormone Receptor and HER2 Status

Subgroup analyses stratified by HR and HER2 status (Figures 4d–g) demonstrated generally robust predictive performance of BC-BioMIXER across molecular subtypes. A modest reduction in AUC was observed in the HER2-positive/HR-positive subgroup relative to the overall cohort, suggesting potential biological heterogeneity that may influence prediction performance in this subgroup.

### Prognostic Value for 3-Year Distant Recurrence-Free Survival

Using a predefined probability threshold of 0.50, patients were dichotomized into predicted pCR and predicted non-pCR groups. In the development cohort (cohort 1), 41.89% of patients were classified as predicted pCR and 58.02% as predicted non-pCR, with similar distributions observed in cohorts 2, 3, and 4. Kaplan–Meier analysis demonstrated significantly superior 3-year DRFS in the predicted pCR group compared with the predicted non-pCR group (HR = 0.36; 95% CI, 0.20–0.67; p < 0.001; Figure 5a). Importantly, among patients who did not achieve pCR at surgery, those predicted by BC-BioMIXER to achieve pCR exhibited significantly improved DRFS (HR = 0.49; 95% CI, 0.25–0.97; p = 0.036; Figure 5c), indicating incremental prognostic value beyond the observed pathological response. No significant DRFS stratification was observed among patients who achieved pCR at surgery.

**Figure 5.**
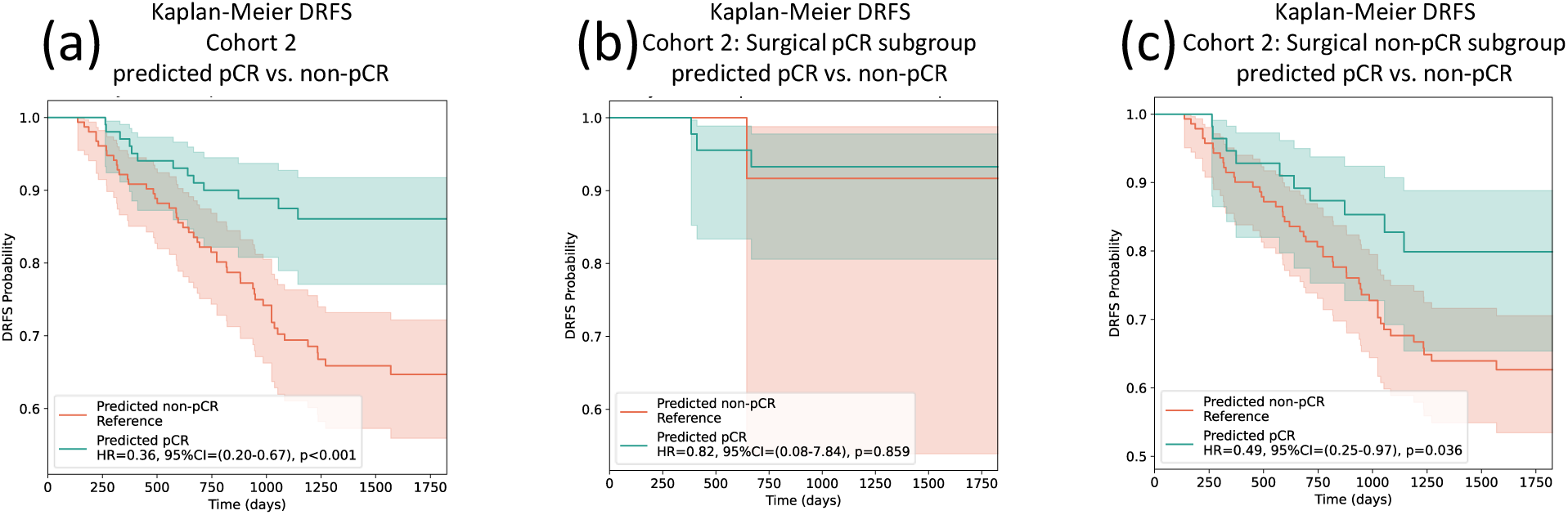
Prognostic value of BC-BioMIXER for DRFS. (a) Kaplan–Meier DRFS estimates for BC-BioMIXER–predicted pCR and non-pCR groups. (b–c) DRFS outcomes stratified by predicted pCR status within the (b) surgical pCR and (c) surgical non-pCR cohorts, demonstrating the model’s ability to refine risk stratification within clinical response groups.

### Personalizing Immunotherapy: Prediction of pCR and Treatment-Specific Outcomes

We further examined the performance of BC-BioMIXER in patients treated with NACT + ICI. In this subgroup, the model maintained robust predictive accuracy, with AUCs of 0.82 (95% CI, 0.73–0.92) in cohort 2 and 0.81 (95% CI, 0.72–0.90) in cohort 4 (Figure 6a). In cohort 2, BC-BioMIXER numerically outperformed CD274 (PD-L1) expression for pCR prediction (AUC 0.82 vs. 0.72), although this difference did not reach statistical signi icance (De ong’s test, p = 0.153).

**Figure 6.**
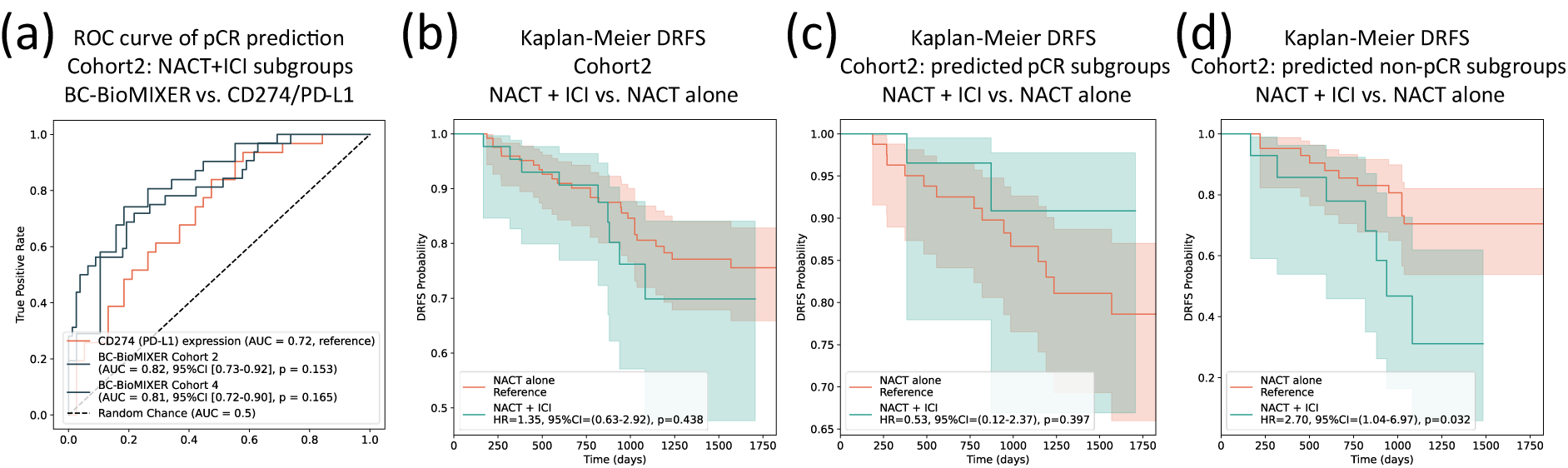
Immunotherapy subgroup analysis and treatment guidance. (a) ROC curves comparing BC-BioMIXER and CD274/PD-L1 expression for pCR prediction. (b) Kaplan–Meier DRFS curves for NACT+ICI vs. NACT alone (total cohort). (c–d) DRFS according to treatment arm within the (c) BC-BioMIXER–predicted pCR subgroup and (d) predicted non-pCR subgroup, highlighting the model’s potential to guide ICI therapy.

When comparing DRFS between treatment arms in cohort 2, no significant difference was observed between patients receiving NACT + ICI and those receiving NACT alone (HR = 1.35; 95% CI, 0.63–2.92; p = 0. 38; Figure 6b). However, within the BC-BioMIXER-predicted non-pCR subgroup, patients treated with ICI experienced significantly inferior DRFS compared with those receiving NACT alone (HR = 2.70; 95% CI, 1.04–6.97; p = 0.032; Figure 6d). In contrast, no significant difference in DRFS was observed between treatment regimens within the predicted pCR subgroup (HR = 0.53; 95% CI, 0.12–2.37; p = 0.397; Figure 6c).

Together, these findings highlight the potential of BC-BioMIXER to support personalized immunotherapy decision-making by identifying patients who may derive limited benefit from additional immune checkpoint inhibition.

## Discussion

This study introduces BC-BioMIXER, a biologically informed predictive framework for NACT response in breast cancer that distills multi-omics knowledge into a model relying exclusively on routinely available clinical data. By integrating molecular insights into imaging- and clinic-based representations, BC-BioMIXER aims to extend the benefits of precision oncology to clinical settings in which comprehensive molecular profiling is unavailable, impractical, or cost-prohibitive. Across both controlled clinical trial cohorts (I-SPY1 and I-SPY2) and an independent, contemporary real-world cohort, BC-BioMIXER achieved pCR prediction performance comparable to a multi-omics reference model and demonstrated robust generalizability despite substantial differences in baseline characteristics and treatment patterns.

From a methodological perspective, this work establishes a generalizable paradigm for transferring biologically meaningful predictive signals from high-dimensional omics data to routine clinical variables. From a clinical standpoint, BC-BioMIXER provides a deployable precision oncology tool for patients undergoing NACT, leveraging only widely accessible inputs—DCE-MRI features and receptor subtype status—thereby enabling biologically informed decision-making in the absence of molecular profiling.

### Model Performance in an Evolving Real-World Treatment Landscape

The clinical relevance of BC-BioMIXER was further supported by its stable performance in a real-world cohort characterized by evolving treatment strategies and patient demographics. Contemporary NACT is increasingly administered to patients with HER2-positive and TNBC, reflecting shifts in therapeutic indications and clinical practice patterns (32). The independently and consecutively collected cohort from Shandong Cancer Hospital captured this transition, exhibiting a markedly higher proportion of HER2-positive and TNBC cases compared with the I-SPY2 trial population.

Despite these differences, BC-BioMIXER maintained consistent predictive performance in this challenging setting. This robustness likely reflects two key design elements. First, explicit incorporation of HR and HER2 status as model covariates enables intrinsic adjustment for major biological drivers of treatment response. Second, the exclusion of low-repeatability latent features during model development promotes reliance on stable, biologically relevant signals derived from routine data, thereby enhancing resilience to cohort heterogeneity and real-world variability (33).

### Clinical Utility: Informing Immunotherapy De-escalation

Beyond response prediction, BC-BioMIXER demonstrated potential utility in informing immunotherapy decision-making. Although the model was developed to predict pathological complete response, it retained stable predictive performance in patients treated with PD-1/PD-L1 inhibitors. More importantly, it enabled identification of a subgroup of patients for whom the addition of ICI to NACT may warrant reconsideration.

Among HER2-negative patients predicted by BC-BioMIXER to be non-responders, those receiving NACT plus ICI exhibited significantly inferior 3-year DRFS compared with those treated with NACT alone. While this observation was derived from retrospective analyses and should be interpreted cautiously, it raises a clinically relevant, hypothesis-generating signal suggesting that a subset of patients—particularly within the HER2-negative and potentially TNBC population—may derive limited benefit from ICI intensification and could experience unfavorable long-term outcomes.

This consideration is especially related given accumulating evidence of increased immune-related adverse events associated with ICI therapy (34). Accurate identification of patients unlikely to benefit from immunotherapy could therefore help preserve treatment efficacy while minimizing unnecessary toxicity.

Notably, this signal is not isolated. Preliminary analyses from the pembrolizumab arm of the I-SPY2 trial have suggested a similar trend toward inferior outcomes among non-pCR patients treated with ICI (35). Our findings extend this observation by demonstrating that such risk stratification may be achievable prior to treatment initiation using routine clinical data. Although large randomized trials such as KEYNOTE-522 have reported improved event-free survival with ICI addition, DRFS outcomes specifically within the non-pCR subgroup remain incompletely characterized. Given the current uncertainty in the literature and the clinically meaningful pattern observed here, continued longitudinal follow-up of real-world cohorts will be essential to clarify the long-term implications of immunotherapy in biologically defined non-responder populations.

### A Knowledge-Transfer Paradigm for Biologically Informed Prediction

Accurate prediction of pCR from routine clinical data was enabled by a novel cross-modal knowledge-transfer paradigm (36). The central premise of this approach is that clinically actionable predictive information is partially shared across modalities—including medical imaging, transcriptomics, and proteomics—and that this shared information can be distilled into representations derived from a more accessible modality.

Previous studies have demonstrated associations between imaging phenotypes and molecular profiles (37), often using imaging as a non-invasive surrogate for genomic or transcriptomic measurements (38) or as a source of novel predictive biomarkers (9). However, comprehensive transfer of integrated multi-omics knowledge into a routinely acquired clinical modality has remained largely unexplored. Leveraging a uniquely rich dataset containing paired transcriptomic, proteomic, imaging, and clinical data (5), our study addresses this gap by explicitly aligning routine imaging-based representations with a biologically informed latent space learned from multi-omics data.

We acknowledge that a routine-data model cannot capture the full informational depth of dedicated molecular profiling and is therefore not expected to outperform a true multi-omics model (6). Nonetheless, this design confers a distinct advantage: predictions are grounded in representations that reflect shared molecular–imaging biology, enhancing biological plausibility and interpretability while preserving clinical feasibility.

### Future Directions

Several avenues warrant further investigation. First, the BC-BioMIXER framework may be extended beyond pCR prediction to model specific metastatic patterns or long-term survival endpoints. Second, incorporation of dynamic biological features, such as longitudinal changes in HER2 status during treatment, may further refine predictive performance. Third, although not formally evaluated in this study, the reliance on routine clinical data suggests potential health-economic advantages by reducing dependence on costly molecular assays. Finally, deeper integration with radiogenomic analyses may further enhance model interpretability by elucidating the biological underpinnings of imaging-derived predictive features.

### Limitations

This study has several limitations. First, the retrospective design introduces the potential for selection bias and unmeasured confounding, particularly in real-world datasets with incomplete data capture. Although statistical strategies were applied to mitigate these effects, prospective validation remains essential. To address this, prospective data collection for external validation is currently underway. Second, the real-world cohort was derived from a single institution, which may limit generalizability despite its contemporary nature. Multi-center validation across diverse healthcare settings will be required to confirm robustness. Third, the most speculative implication of this work—the potential for immunotherapy de-escalation in model-predicted non-responders—should be regarded as hypothesis-generating. Rigorous prospective trials are necessary to establish causal relationships and to determine whether treatment intensity can be safely reduced without compromising oncologic outcomes.

## Conclusion

In conclusion, BC-BioMIXER represents a meaningful advance toward accessible precision oncology for patients receiving neoadjuvant chemotherapy for breast cancer. By translating multi-omics–derived biological insights into a predictive framework based solely on routinely available clinical data, the model achieves robust performance across both clinical trial populations and contemporary real-world settings. Beyond response prediction, BC-BioMIXER highlights a potential role in informing personalized immunotherapy strategies. Collectively, these findings underscore the promise of biologically informed knowledge-transfer approaches to bridge the gap between molecular discovery and scalable clinical implementation.

## Data Availability

All data produced in the present study are available upon reasonable request to the authors

## Supplementary

**Figure S 1.**
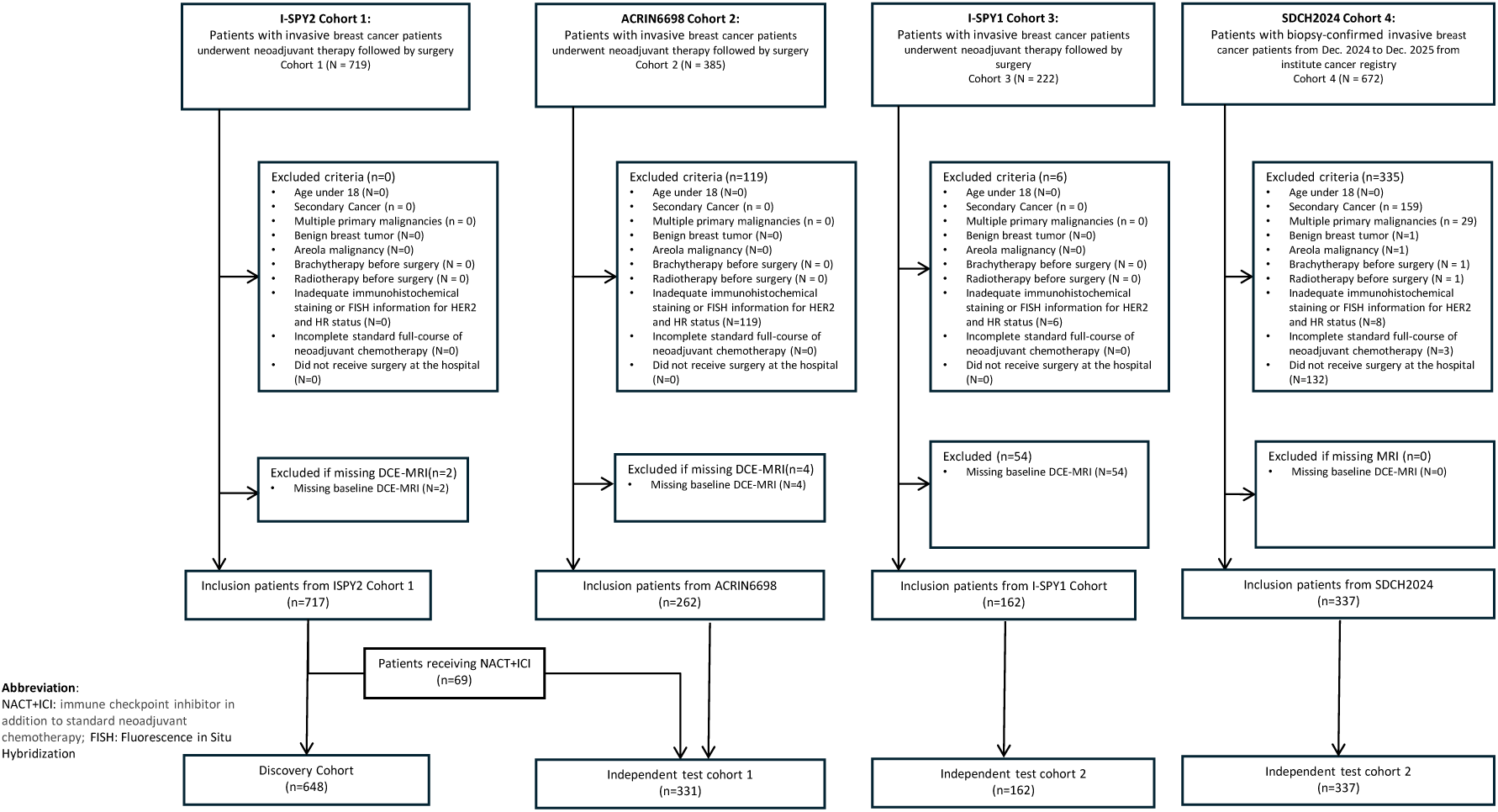
Patient Selection Flowchart of the study.

